# Aberrant flexibility of dynamic brain network in patients with autism spectrum disorder

**DOI:** 10.1101/2024.07.14.24310380

**Authors:** Hui Zhang, Dehong Peng, Shixiong Tang, Anyao Bi, Yicheng Long

**Author notes:** **Correspondence:** Dehong Peng.

## Abstract

**Introduction:** Autism spectrum disorder (ASD) is a collection of neurodevelopmental disorders whose pathobiology remains elusive. This study aimed to investigate the possible neural mechanisms underlying ASD using a dynamic brain network model and a relatively large-sample, multi-site dataset.

**Methods:** Resting-state functional magnetic resonance imaging data were acquired from 208 ASD patients and 227 typical development (TD) controls, who were drawn from the multi-site Autism Brain Imaging Data Exchange (ABIDE)-Preproceesed database. Brain network flexibilities were estimated and compared between the ASD and TD groups at both global and local levels, after adjusting for sex, age, head motion, and site effects. Correlations between the Autism Diagnostic Observation Schedule (ADOS) total score and brain network measures were also investigated after adjusting for the same above covariates.

**Results:** Significantly increased brain network flexibilities (indicating a decreased stability) at the global level, as well as at the local level within the default-mode and sensorimotor areas were found in ASD patients than TD participants. Additionally, significant ASD-related decreases in flexibilities (indicating excessively increased stability) were also observed in several occipital regions at the nodal level. Most of these changes were significantly correlated with the ADOS total score in the entire sample.

**Conclusion:** The results in this study suggested that ASD is characterized by significant changes in temporal stabilities of functional brain network. Our results also pointed to ASD-related dysfunctions in the default-mode, sensorimotor, and occipital systems from a perspective of brain network stability, which can further strengthen our understanding of the pathobiology of ASD.

## 1. Introduction

Autism spectrum disorder (ASD) is a collection of neurodevelopmental disorders which are characterized by early social communication deficits and impaired repetitive behaviors/interests (1,2). The prevalence of ASD is growing in the past decades, leading to high economic burdens globally (1,3). However, the pathobiology of ASD remains elusive; multiple genetic mutations, maternal immune activations, and environmental factors were thought to be involved in the development of ASD (2,4).

Functional magnetic resonance imaging (fMRI) has been widely used in clinical studies, as a non-invasive and convenient method, to investigate the neural mechanisms underlying many common mental disorders (e.g., major depressive disorder and schizophrenia) (5–7). Past fMRI studies have demonstrated that ASD is associated with aberrant brain functions such as significantly decreased/increased functional connectivity (FC) within the visual, frontoparietal (cognition), and language-related subnetworks in the brain (8–11). These studies have significantly improved our understanding of the complex pathobiology of ASD.

The traditional fMRI studies were usually performed under the assumption that FCs within or between different brain networks would be “static (never change)” over time during the fMRI scans. In recent years, however, it was promoted that FCs within/between different brain networks would actually fluctuate over time, and that constructing a “dynamic” rather than “static” brain network model may be valuable to capture important information ignored by traditional brain network models (12–14). Research on properties of “dynamic brain network” under such a framework are emerging, with various measures of temporal variability/stability of the brain networks proposed by different researchers (15–18). Specifically, the “flexibility” (also named “switching rate” by some researchers) of a dynamic functional brain network, which estimates the temporal stability of brain network according to its changing frequencies of modularity structures, has been proved to valid (19) and widely used in recent studies (20–28). In these studies, the flexibility of brain network has been associated with cognition (20,21), emotion (26), aging (24), as well as many common neuropsychiatric disorders such as Parkinson’s disease (27), attention-deficit/hyperactivity disorder (25), schizophrenia (22), anxiety disorder (28), and major depressive disorder (23). Notably, a published study by Harlalka et al. (29) has reported that atypical flexibility of the dynamic functional brain network can quantify (positively associated with) the illness severity in ASD patients. A significantly increased brain network flexibility when compared to matched normal controls has been thought to be reflective of brain dysfunction, which indicates a decreased stability of the brain network organization (30,31). Therefore, the observed positive correlation of symptom severity with flexibility in ASD patients (29) has suggested that a disrupted stability of the brain network may be related to the pathobiology of ASD.

Nevertheless, there are still some notable limitations in the above-mentioned earlier work by Harlalka et al. (29). First, their sample size is relatively small: only 72 patients with ASD and 72 typical development (TD) controls from a single study site was included (29). The previous researchers have suggested that for human connectome studies, satisfactory statistical power and reliability can be achieved when the sample size is larger than a cutoff of approximately 250 subjects (32). Therefore, repeating the study with a larger sample size might be necessary to obtain more reliable conclusion. Second, besides brain network flexibility at the global level, the flexibility can be also estimated at the subnetwork level for particular subsystems of the brain (e.g., for the default-mode, visual, and frontoparietal subnetworks). For instance, using the measure of brain network flexibility, a study by Huang et al. (30) suggested that childhood trauma experiences are linked to decreased temporal stabilities within the default-mode, fronto-parietal, cingulo-opercular, and occipital subnetworks in the brain network. Beyond the global-level brain network flexibility, such results could further strengthen our understanding of the relationship between brain network stability and psychiatric symptoms/disorders at the local level. However, in the earlier study by Harlalka et al. (29), there were no results reported on the possible relationship between brain network flexibility and the ASD at the subnetwork level, which deserves further investigation. Third, when constructing the dynamic functional brain network, an anatomical-based Automated Anatomical Labeling (AAL) atlas with 90 regions of interest (ROIs) was used in the study by Harlalka et al. (29). However, it has been well documented that in studies on human functional brain networks, the anatomical-based AAL atlas would have a poorer performance compared to those higher-resolution functional atlases, such as the Dosenbach atlas (with 160 ROIs) and the Power atlas (with 264 ROIs) (32–34). Thus, performing the analyses based on a functional atlas might be also meaningful for obtaining more accurate results.

To fill the gaps as mentioned above, this study aimed to investigate the possible associations between changes in the brain network flexibility and ASD using a multi-site resting-state fMRI dataset with a relatively large sample size (∼500 participants in total). Specifically, a widely used, validated functional atlas (Dosenbach atlas) was used, and the brain network flexibility was compared between patients with ASD and TD controls at both the global and local (subnetwork) levels. By using a much larger sample and potentially better methodology, we anticipate that the results will provide more reliable and accurate conclusions on the basis of prior work, as well as contribute to a deeper understanding of the pathobiology of ASD.

## 2. Methods and Materials

### 2.1. Participants

The analyzed sample in this study consisted of 435 subjects (208 ASD patients and 227 healthy TD participants) from 9 study sites. Such a sample was drawn from the open-access, multi-site Autism Brain Imaging Data Exchange (ABIDE)-Preproceesed database (http://preprocessed-connectomes-project.org/abide/) (35,36). Details about the participant recruitment, assessment, resting-state fMRI neuroimaging data acquisition, and data preprocessing protocols can be found on the ABIDE website and prior publications (35,36). Most participants completed the Autism Diagnostic Observation Schedule (ADOS) (37) to assess the severity of ASD symptoms.

The original ABIDE-Preprocessed database includes more than one thousand participants recruited from 16 different study sites. In this study, we selected the participants in the analyzed sample based on the following steps: 1) the participants whose demographic information was incomplete (e.g., sex and age) were firstly excluded; 2) participants were excluded when the repetition time (TR) ≠ 2 seconds during the fMRI scans, to prevent biases caused by different temporal resolutions of the dynamic brain networks (38,39); 3) participant with poor fMRI data quality were excluded, as defined by a mean framewise displacement (FD) > 0.2 mm (39,40), bad image coverage (signal loss in any ROI in the Dosenbach atlas), or any of the 3 independent raters gave a “Fail” or “Maybe” rating when performing the manual data checking during the ABIDE preprocessing pipeline (29); 4) finally, the sites with only less than 10 individuals left after the above steps were excluded from the analysis, as did in some other multi-site fMRI studies (40,41). The final analyzed sample of 435 subjects were from the following nine study sites: one from the New York University (NYU), one from the Olin Center (OLIN), one from the San Diego State University (SDSU), one from the Trinity Centre for Health Sciences (TRINITY), one from the University of California Los Angeles (UCLA), two from the University of Miami (UM_1 and UM_2), one from the University of Utah School of Medicine (USM), as well as one from the Yale Child Study Center (YALE) (see **Table 1** for detailed number of participants in each site).

**Table 1.** Number of participants from each site in the analyzed sample.

| Site | ASD | TD | Total |
| --- | --- | --- | --- |
| NYU | 65 | 83 | 148 |
| OLIN | 11 | 6 | 17 |
| SDSU | 8 | 15 | 23 |
| TRINITY | 17 | 19 | 36 |
| UCLA | 20 | 15 | 35 |
| UM_1 | 26 | 43 | 69 |
| UM_2 | 12 | 18 | 30 |
| USM | 32 | 18 | 50 |
| YALE | 17 | 10 | 27 |
| Total | 208 | 227 | 435 |

Abbreviations: ASD = autism spectrum disorder; TD = typical development; the full names of each site can be found in the main text of the **Section 2.1**.

The ABIDE-Preprocessed data is open shared, and Institutional Review Board (IRB) approval was provided by each site (data contributor) in the ABIDE database.

### 2.2. Neuroimaging data acquisition and preprocessing

The resting-state fMRI data downloaded from the ABIDE database has been preprocessed using the pipelines whose details can be found at: http://preprocessed-connectomes-project.org/abide/Pipelines.html. For all participants, the fMRI scans were acquired and preprocessed in each study site independently. The scans were acquired using SIEMENS, PHILIPS, or GE magnetic resonance imaging machines; the TR was 2 seconds for all scans but there might be differences in other scanning parameters (e.g., echo time and total scan duration). The preprocessing pipeline includes slice timing correction, motion realignment, nuisance signals (motion and tissue signals) removal, and registration. Notably, the ABIDE database provided both the preprocessed data with and without the step of global signal regression (GSR). In the current study, we used the data without GSR since the usage of GSR is still controversial (42,43). Furthermore, to control for confounding effects caused by differences in data acquisition and preprocessing procedures between different site, we included “site” as covariates (using dummy coding) in all analyses (see details in the followed sections).

### 2.3. Construction of dynamic brain networks

The steps of constructing dynamic brain networks and estimating brain network flexibility are summarized in **Figure 1**. A dynamic functional brain network is comprised of a set of nodes (ROIs) and connections (FCs) between nodes, where the strengths of connections change over time (44). In this study, nodes in the dynamic brain network were defined using the 160 ROIs from the Dosenbach atlas (45), which has been validated and widely used in clinical studies (30,46–48). For each participant, the mean time series were extracted from each ROI, and then divided into a number of partly overlapping time windows using the “sliding-window” method which is commonly used in dynamic brain network studies (**Figure 1A**) (49–51). Here, a fixed window width of 100 seconds (50 TRs) and a sliding step length of 2 seconds (1 TR) were used, according to the recommendations in previous works to balance the reliability of results and computational complexity (30,33,39,52). Within each time window, the FC strengths between all possible pairs of ROIs were computed using Pearson correlations, resulting in a “snapshot” of the brain network organization as shown by a 160×160 FC matrix. These time-ordered “snapshots” (matrices) then formed a multilayer dynamic brain network *G* = (*G_t_*)*_t = 1, 2, 3, …, T_*, where the *t*th “snapshot”/matrix (Gt) represents the brain network organization at the *t*th time window (**Figure 1B**). Note that the total number of time windows (*T*) was determined by the length of fMRI scanning and was thus different between different study sites.

**Figure 1.**
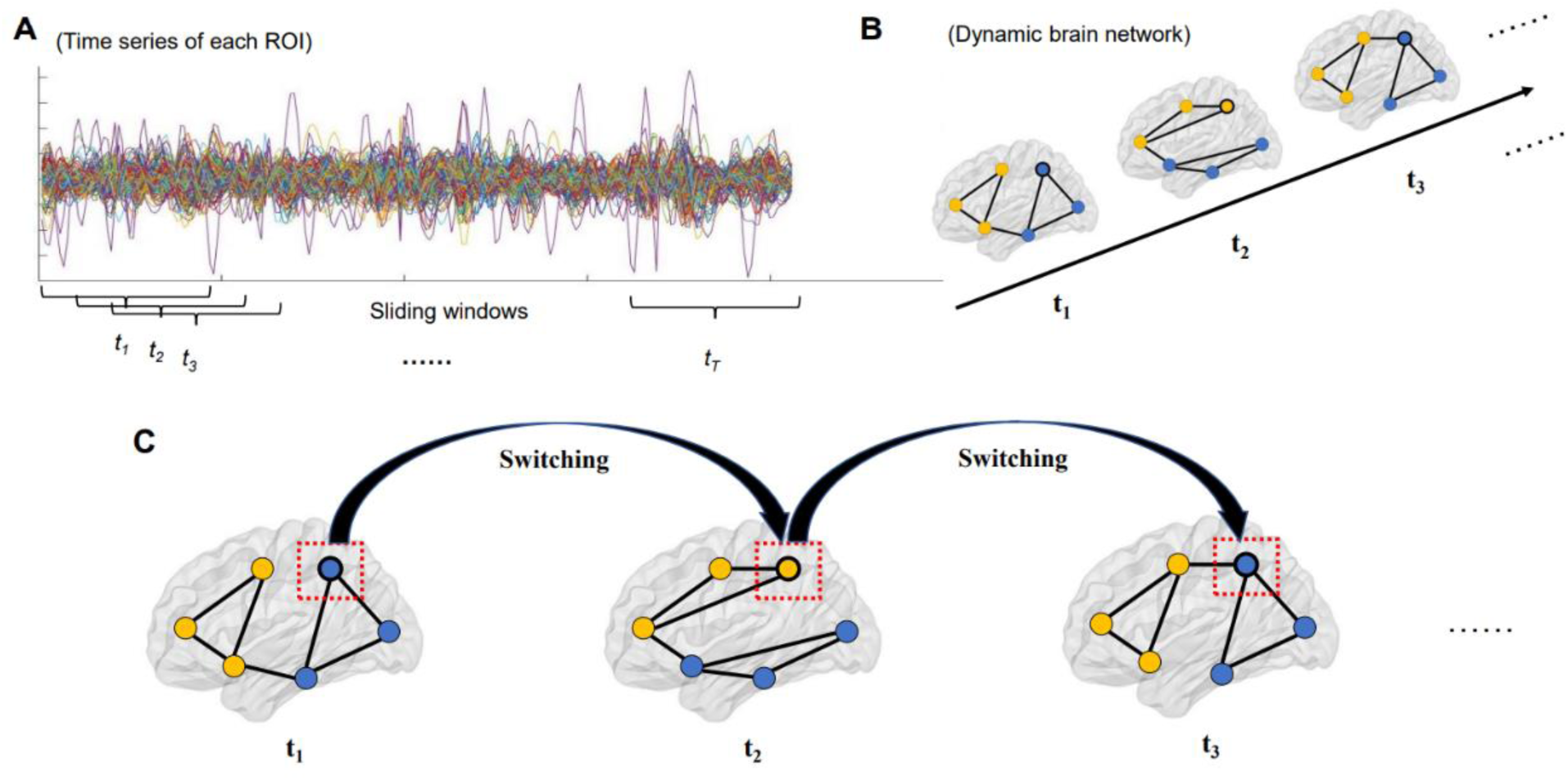
Steps of constructing dynamic brain networks and calculating flexibility (see details in **Sections 2.3** and **2.4**). (**A**) The time series of each region of interest (ROI) were divided into a number of time windows using the sliding-windows approach. (**B**) Brain network organizations were constructed for each time window, which formed a dynamic brain network. (**C**) Flexibilities (switching rates) of each ROI were then estimated by number of times for which it switched from one “community” to another.

### 2.4. Estimating brain network flexibility

After constructing the dynamic brain network for each participant, the brain network flexibility was then computed strictly following some previous publications (20,30,47,53–55). Briefly, a dynamic community detection algorithm as described by Mucha et al. (54) was implemented in Matlab using an open-source code package at: https://github.com/GenLouvain/GenLouvain (56). Based on such an algorithm, all nodes (ROIs) in the dynamic brain network were assigned into several communities at each time window, and result in different community assignments for different time windows. The “flexibility/switching rate” of a node *i* (*f_i_*) can be then computed based on its switching frequency between different communities over time as:

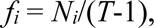

where *N_i_* is the number of times for which the node *i* “switched” from one community to another (**Figure 1C**) (57). The calculation was performed with the assistance of the Network Community Toolbox (http://commdetect.weebly.com/) (57). Notably, since individual runs of the algorithm could lead to slightly different community assignments, the algorithm and flexibility calculation were repeated for a total of 100 times, and the final flexibility values were averaged across the 100 runs (30,47,55,58). After that, flexibility of the whole brain network (global-level flexibility) was obtained by averaging flexibilities of all the 160 ROIs.

According to previous research (45), all ROIs from the Dosenbach atlas can be assigned into six subnetworks including the default-mode, occipital, cingulo-opercular, fronto-parietal, sensorimotor, and cerebellar subnetworks. On the basis of such an assignment, flexibilities for each of the six subnetworks were further obtained by averaging all nodes belonging to each particular subnetwork (22,30,47).

### 2.5. Statistics

Demographic and clinical characteristics were compared between the ASD and TD groups using two-sample *t* tests or Chi-square tests as appropriate. All brain network measures (flexibilities at the global, subnetwork, and nodal levels) were compared between the ASD and TD groups by the analysis of covariance (ANCOVA) covarying for age, sex, head motion (mean FD), and site (dummy coded). False discovery rate (FDR) corrections were performed to correct for multiple comparisons across the six subnetworks or across the 160 nodes. Significance was set at a FDR-corrected *p* < 0.05.

When significant between-group differences were found on any brain network measures, post-hoc correlation tests were further performed to investigate their possible relationships with the severity of clinical symptoms (measure by the total ADOS score) in the ASD patients. Here, partial Pearson correlations adjusted for age, sex, head motion and site effects were performed between the brain network measures and ADOS total score. The correlation analyses were performed in the entire sample and in the ASD group independently, respectively. Similar to the between-group comparisons, FDR corrections were performed to correct for multiple correlation tests across multiple subnetworks or nodes. Significance was set at a FDR-corrected *p* < 0.05, too.

## 3. Results

### 3.1. Sample characteristics

Demographic and clinical characteristics of the ASD and TD groups are presented in **Table 2**. There was no significant group difference in age (*t* = 0.847, *p* = 0.398); nevertheless, the ASD group had a significantly higher proportion of males and a higher mean FD value than the TD group (both *p* < 0.05). Furthermore, the ASD group had a significantly higher mean ADOS total score than the TD group (*t* = 20.385, *p* < 0.001), which is not surprising.

**Table 2.** Comparisons on demographic and clinical characteristics between the ASD and TD groups.

|  | ASD (*n* = 208),<br>mean ± SD | TD (*n* = 227),<br>mean ± SD | Group comparisons |
| --- | --- | --- | --- |
| Age (years) | 16.35 ± 6.72 | 15.86 ± 5.16 | *t* = 0.847, *p* = 0.398 |
| Sex (male/female) | 179/29 | 171/56 | $\chi^2 = 7.945$, *p* = 0.005 |
| Mean FD (mm) | 0.09 ± 0.05 | 0.06 ± 0.03 | *t* = 5.715, *p* < 0.001 |
| AODS total score<sup>a</sup> | 8.48 ± 5.90 | 0.08 ± 0.44 | *t* = 20.385, *p* < 0.001 |

Abbreviations: AODS = the Autism Diagnostic Observation Schedule; ASD = autism spectrum disorder; FD = framewise displacement; TD = typical development.
<sup>a</sup>Data on the AODS scores were available for 206 ASD patients and 225 TD participants, respectively.

### 3.2. Group comparisons on flexibilities

Compared to the TD group, the ASD patients showed a significantly higher brain network flexibility at the global level (*F* = 4.280, *p* = 0.039) (**Figure 2A**). Furthermore, at the subnetwork level, the ASD patients showed significantly higher flexibilities in the default-mode and sensorimotor subnetworks than TD participants (*F* = 11.404/7.730, FDR-corrected *p* = 0.006/0.018 for the default-mode/sensorimotor subnetworks, respectively) (**Figure 2B**). Compared to the TD group, significant higher nodal level flexibilities in ASD patients were found in several ROIs belonging to the default-mode subnetwork, including the precuneus, angular gyrus, and post cingulate cortex (all corrected-*p* < 0.05, as marked in red in **Figure 3**); meanwhile, significant lower nodal level flexibilities in ASD patients were found in two ROIs within the occipital subnetwork, which are located in the occipital and post-occipital regions, respectively (both corrected-*p* < 0.05, as marked in yellow in **Figure 3**). More details about the nodes showing significant between-group differences can be found in **Supplementary Table S1**.

**Figure 2.**
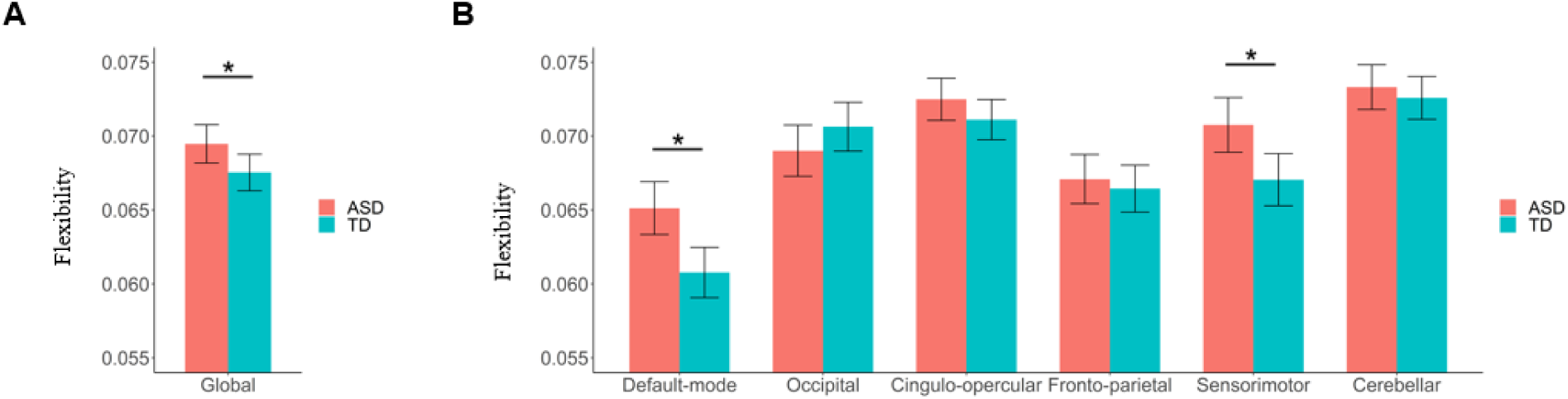
Results of comparisons on global and subnetwork-level brain network flexibilities between the autism spectrum disorder (ASD) and typical development (TD) groups. (**A**) Comparison on flexibility at the global level. (**B**) Comparisons on flexibilities at the subnetwork level. The “*” indicates a significant between-group difference with corrected-*p* < 0.05.

**Figure 3.**
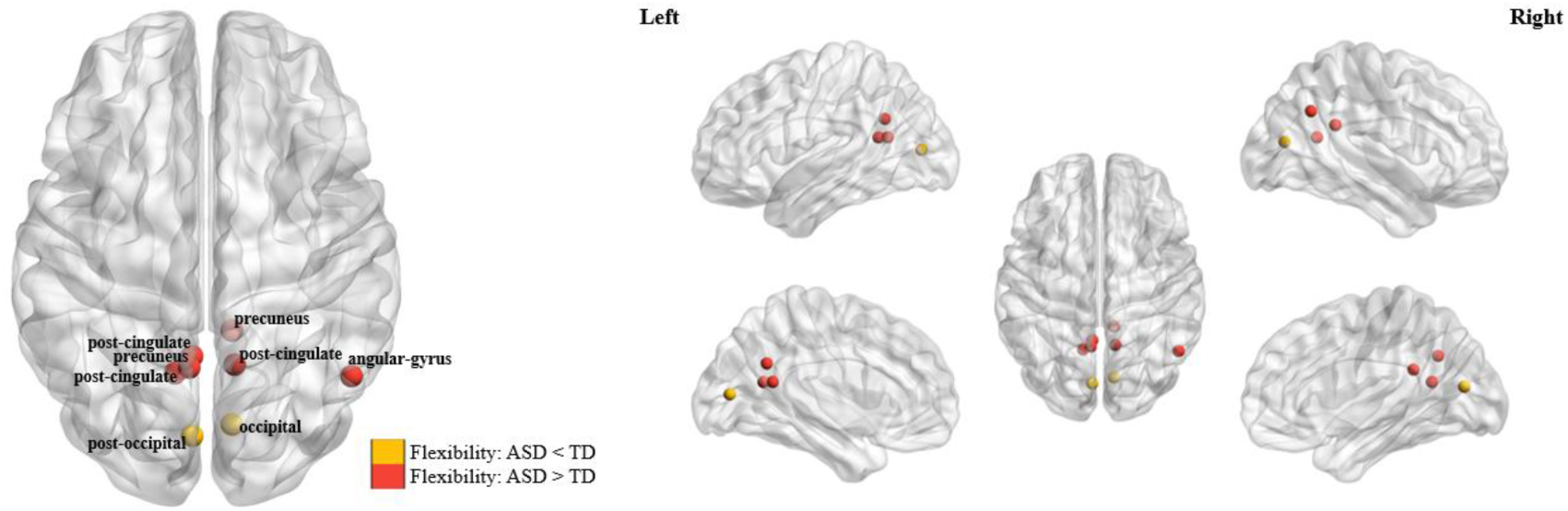
The brain nodes which showed significant differences in flexibility (corrected-*p* < 0.05) between the autism spectrum disorder (ASD) and typical development (TD) groups.

### 3.3. Correlation analyses

When testing correlations in the entire sample, the AODS total score was found to be significantly positively associated with flexibilities in the default-mode and sensorimotor subnetworks flexibility, after adjusting for sex, age, head motion, and site effects (*r* = 0.109/0.134, FDR-corrected *p* = 0.024/0.010 for the default-mode/sensorimotor subnetworks, respectively) (**Figure 4**). Moreover, the AODS total score was found to be significantly associated with flexibilities in several nodes belonging to the default-mode and occipital subnetworks (**Supplementary Table S2**). Nevertheless, no significant results were found when correlations were tested in the ASD group independently (corrected *p* > 0.05).

**Figure 4.**
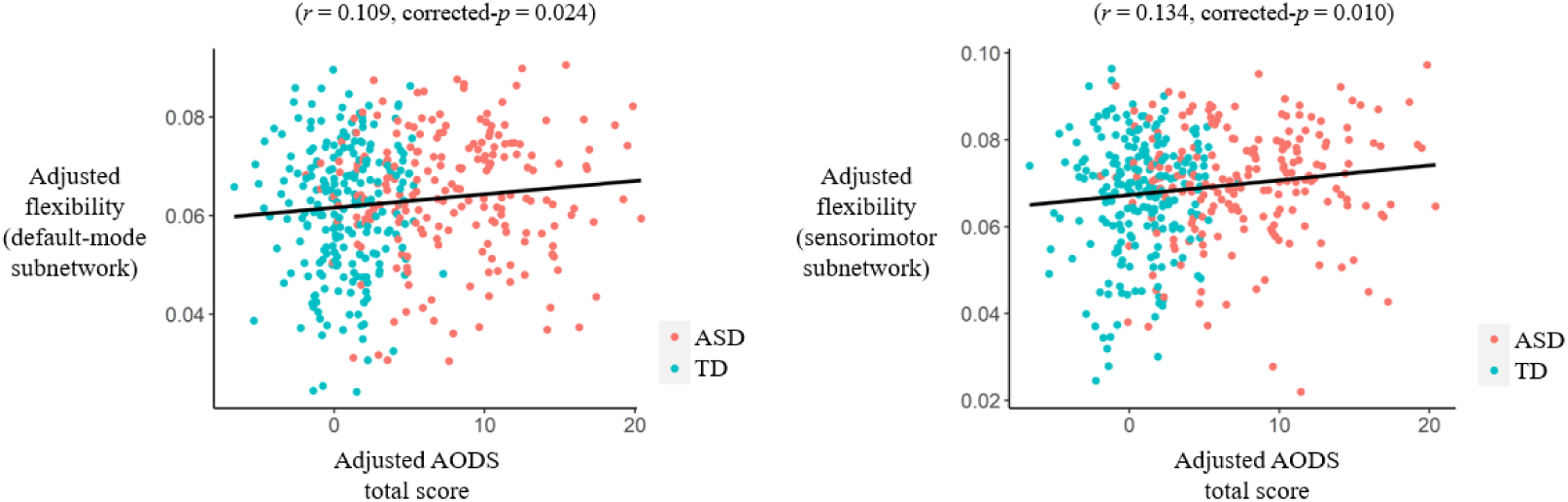
Results of partial correlations (adjusted for sex, age, head motion, and site effects) between the Autism Diagnostic Observation Schedule (AODS) total score and subnetwork-level flexibilities in the entire sample. ASD = autism spectrum disorder; TD = typical development.

## 4. Discussion

In the present study, we explored the possible neural mechanisms underlying ASD using a measure of “flexibility” based on the dynamic brain network model, and a relatively large-sample, multi-site dataset. When compared to TD participants, the most significant findings in ASD patients included a higher brain network flexibility at the global level, as well as higher flexibilities within the default-mode/sensorimotor areas and lower flexibilities within the occipital areas at the local levels. These findings may strengthen our understanding of the pathobiology of ASD from the perspective of brain network stability.

At the global level, we observed a significantly higher flexibility (switching rate) in the ASD patients than TD controls (**Figure 2A**), which is in line with an earlier study with relatively small sample size by Harlalka et al. (29). It was thought that the human brain network needs to be “flexible” (changing the community structures over time) for cognitive and affective processes; however, an observed excessively increased flexibility than normal controls may also be abnormal, indicating a decreased temporal stability of the brain functional organizations (26,31,53,57). Therefore, our results could further support the opinion by Harlalka et al. that ASD is accompanied by decreased stability in the brain network (29). Some previous studies have reported both decreased and increased temporal stabilities of functional brain network in ASD patients based on other measures (other than flexibility) under a framework of dynamic brain network. For example, using resting-state fMRI data collected from 62 ASD patients/57 TD participants and the “transient connectivity patterns/states”-based analyses, Mash et al. (59) found an increased variability over time (decreased stability) of brain network in ASD patients, which is in line with the results in the current study. However, using fMRI data from 24 ASD patients and 26 TD individuals, an opposite conclusion was reported by Watanabe et al. (60) that ASD is associated with “overly stable neural dynamics” (increased stability) in functional brain network. Notably, by analyzing a multi-site dataset, the sample size in the current study is much larger when compared to most of the above published studies, which could lead to more reliable results (32). Therefore, our results may offer more solid evidence to demonstrate that ASD is associated with a “less stable” functional brain network.

At the local (subnetwork and nodal) levels, it was observed that ASD-related decreases in stabilities of the brain network (increases in brain network flexibilities) were mainly found in the default-mode and sensorimotor areas (**Figure 2B** and **Figure 3**). The default-mode subnetwork in the brain is known to mediate one’s self-referential and internally-directed processing (61), and a decreased stability of it has been linked to multiple common mental illnesses such as depression (39,62). The sensorimotor areas in the brain play important roles in sensorimotor control (63), whose dysfunctions have also been associated with multiple illnesses such as depression and schizophrenia (17,64). Actually, the key functions of these brain systems are well known to be disrupted in ASD supported by previous studies. For example, there has been ample evidence that ASD patients are impaired in one of the key functions of the default-mode subnetwork: self-referential cognition, which is the ability to process social information relative to oneself, and such impairments may be closely related to the social cognitive dysfunction in ASD (65–68). On the other hand, it has been widely reported that sensorimotor skills are atypical in individuals with ASD, and such deficits are associated with severity of ASD symptoms (69–71). Therefore, results of the current study may point to ASD-related dysfunctions in the default-mode and sensorimotor systems from a perspective of brain functional dynamics, which may underlie the clinical features of ASD such as impaired self-referential cognition and sensorimotor difficulties.

At the nodal level, significant decreases in flexibilities (increased stability) in ASD patients were additionally observed in several occipital regions (**Figure 3**). Such alterations, interestingly, are opposite to the observed ASD-related changes at the global level, as well as the changes in other subnetworks. While a decreased stability may suggest abnormal changes in the brain network as mentioned earlier, an excessively “decreased variability” (increased stability) may be also indicative of brain dysfunctions and decreased ability to adapt to changing environmental demands (31,53). The occipital cortex is known to involved in the visual processing of the brain (53,72); previous research has reported significant ASD-related alterations in both structures and functions in the occipital cortex, which were associated with visual processing and social communication deficits in ASD patients (73,74). Our results thus again highlighted the critical importance of focusing on the occipital areas in research on ASD. Furthermore, combining such results with finding in the default-mode/sensorimotor areas, it might be concluded that both excessively increased and decreased functional stabilities (in different brain systems) are involved in the pathobiology of ASD.

Our study has some limitations. First, although positive correlations were shown between the brain network flexibilities and ASD symptom severity in the entire sample (**Figure 4**), no significant results were obtained when testing the correlations in the ASD group independently to support our clinical symptoms-related hypotheses. Second, the current study used cross-sectional data and thus we were unable to ascertain the participants’ developmental trajectories; future studies will benefit from longitudinal designs.

In conclusion, this study investigated the possible relationships between ASD and changes in functional brain network dynamics using a multi-site dataset. The results suggested significantly increased brain network flexibilities (indicating a decreased stability) at the global level, as well as at the local level within the default-mode and sensorimotor areas in ASD patients than TD participants. Additionally, significant ASD-related decreases in flexibilities (indicating excessively increased stability) were also observed in several occipital regions at the nodal level. These results also pointed to ASD-related dysfunctions in the default-mode, sensorimotor, and occipital systems from a perspective of brain network stability, which can further strengthen our understanding of the pathobiology of ASD.

## Data Availability

All data produced in the present study are available upon reasonable request to the authors.

## Author contributions

HZ, ST and YL conceived the idea and structured the manuscript. YL performed the data analysis. HZ and YL drafted the manuscript. DP, ST, and AB revised the manuscript. All authors have read and agreed to the published version of the manuscript.

## Funding

This research was funded by the Natural Science Foundation of Hunan Province, China (2023JJ60438 to Hui Zhang), the Scientific Research Launch Project for new employees of the Second Xiangya Hospital of Central South University (to Bian Yao and Yicheng Long), the Health Research Project of Hunan Provincial Health Commission (W20243225 to Yicheng Long), and the National Natural Science Foundation of China (82201692 to Yicheng Long).

## Conflict of Interest Statement

The authors declare no conflict of interest.

## Notes

### Competing Interest Statement

The authors have declared no competing interest.

### Author Declarations

The study used (or will use) ONLY openly available human data from the Autism Brain Imaging Data Exchange (ABIDE)-Preproceesed database that were originally located at: http://preprocessed-connectomes-project.org/abide/index.html.

